# Reduced expression of N-methyl-D-aspartate receptor and calcium signaling genes in gray matter is associated with cognitive function in patients with breast cancer

**DOI:** 10.64898/2025.12.15.25342306

**Authors:** Shelli R. Kesler, Oscar Y. Franco-Rocha, Manuela Kogon, Sarah Braun, Leah Tolby, Ruth Nyagaka, Alexa De La Torre Schutz, Douglas W. Blayney, Oxana Palesh

## Abstract

Cognitive decline is common after cancer, but little is known regarding the etiology of this adverse effect, especially in terms of molecular mechanisms. This prospective study obtained brain imaging and cognitive testing from 50 newly diagnosed women with primary breast cancer prior to any cancer treatment and 53 female controls. Participants completed up to 7 assessments for a total time span of 9.7 +/- 0.92 years. Imaging transcriptomics was used to measure the expression of genes in the brain involved in N-methyl-D-aspartate (NMDA) and calcium-mediated neurotransmission. *GRIN2A, GRIN2B, CACNA1C* were significantly expressed in gray matter in both groups (R^2^ > 0.094, p < 0.015). *GRIN2A* (t = -2.72, p = 0.007) and *CACNA1C* (t = -2.11, p = 0.036) were significantly lower in the cancer group compared to controls across timepoints. *GRIN2A* declined over time in patients, and this was significantly different compared to controls (χ² = 9.73, p = 0.001). Cognitive scores were significantly lower in patients compared to controls (p = 0.002). In patients, *GRIN2A* were significantly associated with cognitive performance over time (p < 0.007). These findings suggest that gene expression involved in neurotransmission is disrupted in the brain among patients with breast cancer and may contribute to cognitive changes. Our results provide novel molecular insights regarding the roles of non-CNS cancer pathology and treatments in the brain related to calcium signaling and pro-survival/plasticity-related pathways. Our findings also point to potential treatments for cognitive effects of cancer.

## Introduction

Cancer-related cognitive impairment (CRCI, known colloquially as “chemobrain”) is one of the most prevalent and burdensome symptoms, affecting up to 84.4% of survivors.[1] CRCI has been associated with diminished quality of life and increased cancer mortality.[2–4] Notably, survivors have reported cognitive difficulties up to 20-years post-treatment.[5, 6] Despite growing recognition of CRCI, its etiologies remain unclear.

Inflammation, neurotoxic injury, suppression of neuroprogenitor cells, and accelerated aging are among the candidate mechanisms.[7, 8] However, studies focused on molecular mechanisms have been limited. Challenges include the difficulty in translating findings from preclinical models and the incomplete correspondence of peripheral assays with the central nervous system environment.

Imaging transcriptomics is a non-invasive, in vivo method for examining how gene expression differs across brain regions, revealing molecular features linked to disease-related changes in brain structure and function.[9–12] Unlike single nucleotide polymorphism (SNP) studies, imaging transcriptomics offers more direct and functional insights by linking gene expression to brain organization. CRCI is consistently associated with abnormalities in large-scale brain networks that support integrated cognitive functions such as executive control and memory.[13–15] These networks uniquely rely on glutamate receptor and calcium-mediated neurotransmission. N-methyl-D-aspartate receptors (NMDARs) are glutamate receptors that flux high levels of calcium to support the persistent neuronal firing required to maintain advanced cognition, such as executive function.[16] Calcium drives cyclic adenosine 3′,5′-monophosphate production in feedforward signaling cascade that is essential for synaptic plasticity and transcription.[17]

*GRIN2A* and *GRIN2B* genes encode NMDAR -GluN2A and -GluN2B subunits, respectively.[16] *CACNA1C* encodes CAv1.2 calcium channels and cAMP-activating proteins.[18] Dysregulated activation of NMDAR/calcium signaling pathways are associated with various cognitive disorders and neurodegenerative diseases.[16, 19, 20] Abnormal calcium signaling is also associated with increased neuroinflammation,[16, 21] which is putatively elevated in CRCI.[22] In our unique, 10-year prospective, longitudinal study, we measured *GRIN2A, GRIN2B, and CACNA1C* expression in gray matter. We hypothesized that the expression of these genes across time would significantly differ in patients compared to non-cancer controls and would affect cognitive function. Given the novelty of these analyses and the complex interplay of biological systems under investigation, we explored nonlinear relationships and did not make *a priori* predictions regarding direction.

## Methods

### Participants

We prospectively enrolled 50 females newly diagnosed with primary breast cancer (stages 0 to IIIA) prior to any cancer treatment (surgery with anesthesia, chemotherapy, radiation) and 53 noncancer female controls. Participants completed up to 7 different assessments for a total time span of 9.7 +/-0.92 years, on average (Table 1). In the first phase of the study, 3 assessments occurred: at enrollment, 1 month after completing adjuvant chemotherapy treatment (or yoked interval for the control group), and again 1 year later. During the second phase of the study, we annually re-evaluated a subgroup of participants (N=35 per group) for the duration of the project, resulting in an additional 4 assessments (Table 2). Participants were recruited through physician referrals and community-based advertisements. Participants were excluded for history of chronic health conditions affecting cognition, major sensory deficits, or limited English proficiency. Demographic and medical history data were obtained from self-report. This study received Stanford University Institutional Review Board approval, followed Declaration of Helsinki ethics, and obtained written informed consent from all participants.

**Table 1.**
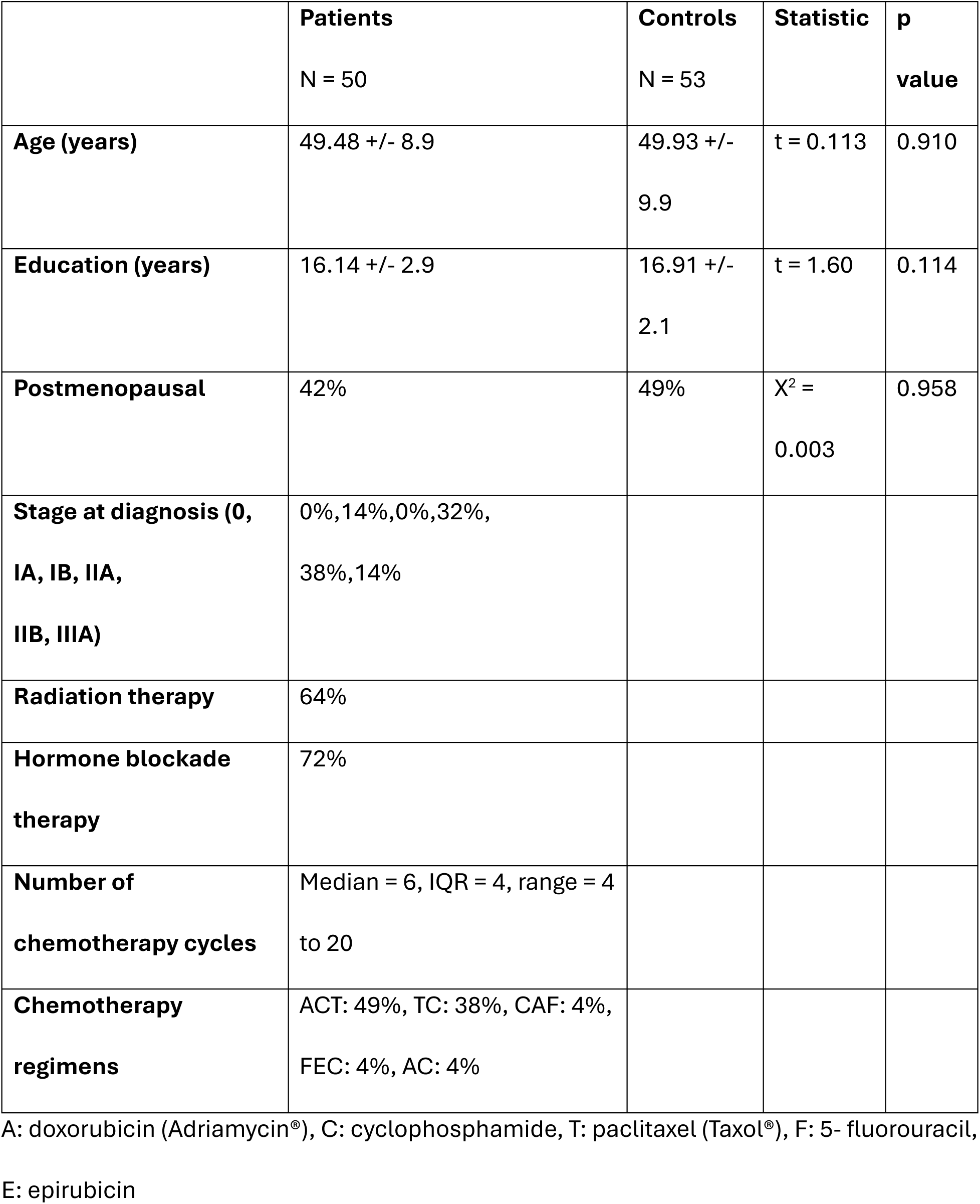
Demographic data at enrollment.

**Table 2.** Number of participants at each assessment. The time in months represents the average interval between the assessment and baseline. There was a longer interval between the third and fourth assessments than planned due to the COVID-19 pandemic which then required a slight acceleration of the final two assessments. The longer interval between the fourth and fifth assessments was due to relocations of the principal investigators.

| **Timepoint** | **1** | **2** | **3** | **4** | **5** | **6** | **7** |
| --- | --- | --- | --- | --- | --- | --- | --- |
| **Time in months** | 0 | 6 | 18 | 75 | 99 | 109 | 117 |
| **Chemo** | 50 | 36 | 42 | 28 | 22 | 17 | 9 |
| **Control** | 53 | 44 | 44 | 37 | 31 | 26 | 19 |

### Imaging Transcriptomics

A high-resolution, anatomic brain MRI scan was obtained for each participant: TR=8.5, TE= minimum, TI=400 ms, FOV=22 cm, slice thickness=1.5 mm, 124 slices, 256 × 256, scan time=4:33 min using a GE Signa HDx whole body scanner (GE Medical Systems, Milwaukee, WI). Additional scans were collected that are not reported here.

We extracted gray matter volumes from the MRI for each participant using voxel-based morphometry.[23] We obtained brain transcriptome data from the Allen Human Brain Atlas (AHBA).[9] The gray matter volume was resampled into AHBA coordinates with a 5mm resolution. The expression data for each of the six AHBA donors, for each gene, was sampled from each of 169 AHBA regions. Principal Components Analysis was performed on the region-by-gene expression matrix to identify components explaining at least 95% variance across the AHBA donors. The component scores were then entered as independent variables into a multiple linear regression with the spatially corresponding gray matter volumes as dependent variable.

The accuracy of each AHBA probe-to-gene mapping was verified and probe data were normalized to z-scores. Representative probes were then selected in a data-driven manner considering between-donor homogeneity and the distributions of probe data.[24] Gene expression reliability across donors was measured using an intraclass correlation coefficient (ICC).

### Cognitive Assessment

We administered Trails 1 and 5, referred to as Trails A and B, of the Comprehensive Trail Making Test. These tests measure processing speed, attention, and executive function.[25] Other cognitive tests were also administered but we *a priori* selected Trails A and B for this study to reduce comparisons and to focus on executive function. Executive function is one of the most profoundly affected cognitive domains in CRCI, and impairments in this area are among the most debilitating.[7, 26]

### Data Analysis

Data were visualized for normality and outliers. Missing data were assessed using Little’s MCAR test and visualizations, which indicated missing at random. We fit generalized additive models (GAMs) using penalized regression splines, with smoothing parameters estimated by restricted maximum likelihood. GAMs are well-suited for irregular or sparse longitudinal data.[27–29] They stabilize estimates at timepoints with fewer observations by borrowing strength from neighboring values through smooth basis functions. The distributions and trajectories of the brain transcriptome are unknown. Thus, we modeled potentially non-linear changes over time, using spline functions with a restricted basis dimension (k = 5) to limit the complexity of the smooth and prevent overfitting in sparsely sampled timepoints. Models included a random intercept as well as covariates for age, education, and gray matter volume.

We fit separate smooth functions of time for each group, allowing non-linear outcome trajectories to vary flexibly between groups. To determine group-by-time effect, we refit the model using maximum likelihood and compared it to a model that included a shared smooth for time to test a group-by-time interaction effect using a likelihood ratio.

To determine the effects of gene expression on cognitive performance, we fit GAMs predicting Trails A/B score as a function of genes that showed significant main effects of group or group by time effects, with age and education as covariates. These GAMs were fit only in patients. All analyses were performed in the R Statistical Package (v4.5.1) with p < 0.05.

## Results

As shown in Table 3, *GRIN2A/B* and *CACNA1C* were significantly expressed in gray matter in both groups (R^2^ > 0.094, p < 0.015). Gene expression showed high reliability (ICC > 0.829).

**Table 3.** Correlation of gene expression with gray matter volume across participants.

|  | **Cancer** |  |  | **Control** |  |  |
| --- | --- | --- | --- | --- | --- | --- |
| **Gene** | *GRIN2A* | *GRIN2B* | *CACNA1C* | *GRIN2A* | *GRIN2B* | *CACNA1C* |
| **R<sup>2</sup>** | 0.377 | 0.094 | 0.477 | 0.372 | 0.092 | 0.476 |
| **F** | 26.01 | 4.45 | 15.13 | 25.45 | 4.38 | 15.08 |
| **p** | < 0.000 | 0.014 | < 0.000 | < 0.000 | 0.015 | < 0.000 |
| **ICC\*** | 0.941 | 0.956 | 0.829 |  |  |  |
| **ICC SD** | 0.014 | 0.013 | 0.043 |  |  |  |

ICC: intraclass correlation coefficient; SD: standard deviation.
\* ICC values are computed across Allen Human Brain Atlas donors (i.e., gene expression reliability), not for sample groups.

*GRIN2A* (t = -2.72, p = 0.007) and *CACNA1C* (t = -2.11, p = 0.036) were significantly lower on average in the cancer group compared to controls across timepoints (Figure 1). *GRIN2A* declined over time in patients, and this was significantly different compared to controls (χ² = 9.73, p = 0.001). Change in *GRIN2B* (χ² = 0.00, p = 1) and *CACNA1C* (χ² = 0.973, p = 0.314) were not different between groups. Age and gray matter volume contributed significantly to all models (F > 3.72, p < 0.013). Higher education was associated with increased *GRIN2A/B* expression (p < 0.047).

**Figure 1.**
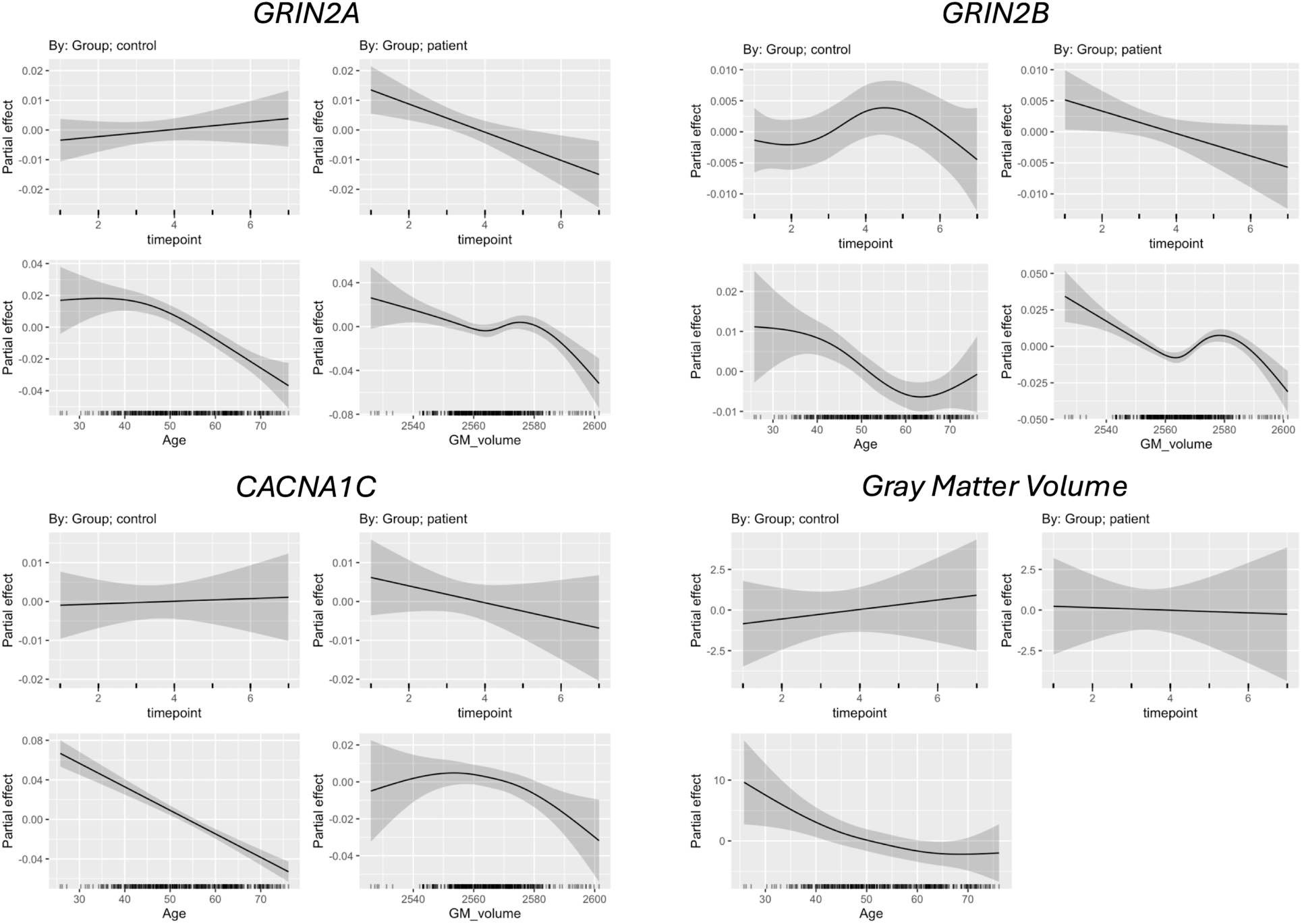
Longitudinal trajectory of neurotransmission-related genes in gray matter and gray matter volume. Expression of *GRIN2A* was significantly lower (t = -2.72, p = 0.007) and declined significantly over time (χ² = 9.73, p = 0.001) in patients compared to controls. *GRIN2B* was not different between groups and did not change significantly over time. *CACNA1C* was significantly lower in patients compared to controls (t = -2.11, p = 0.036) but longitudinal trajectories did not differ between groups. Total gray matter volume was not different between groups (t = -1.41, p = 0.159) and did not change significantly over time. Plots show the partial effects controlling for all other predictors in the model. The solid line indicates the estimated smooth function for the generalized additive model with the shaded area representing the 95% confidence interval.

Gray matter volume was lower in patients but did not differ significantly between groups (t = -1.41, p = 0.159) nor did it change significantly over time in either group (p > 0.529, Figure 1).

Trails A (t = -4.29, p < 0.001) and Trails B (t = -3.14, p = 0.002) scores were significantly lower in patients compared to controls across timepoints (Figure 2). Performance trajectories did not differ between groups (χ² < 0.157, p > 0.695). Higher education was associated with higher performance in both models (p < 0.003) and age contributed to the Trails A model in a nonlinear manner (F = 5.89, p = 0.001).

**Figure 2.**
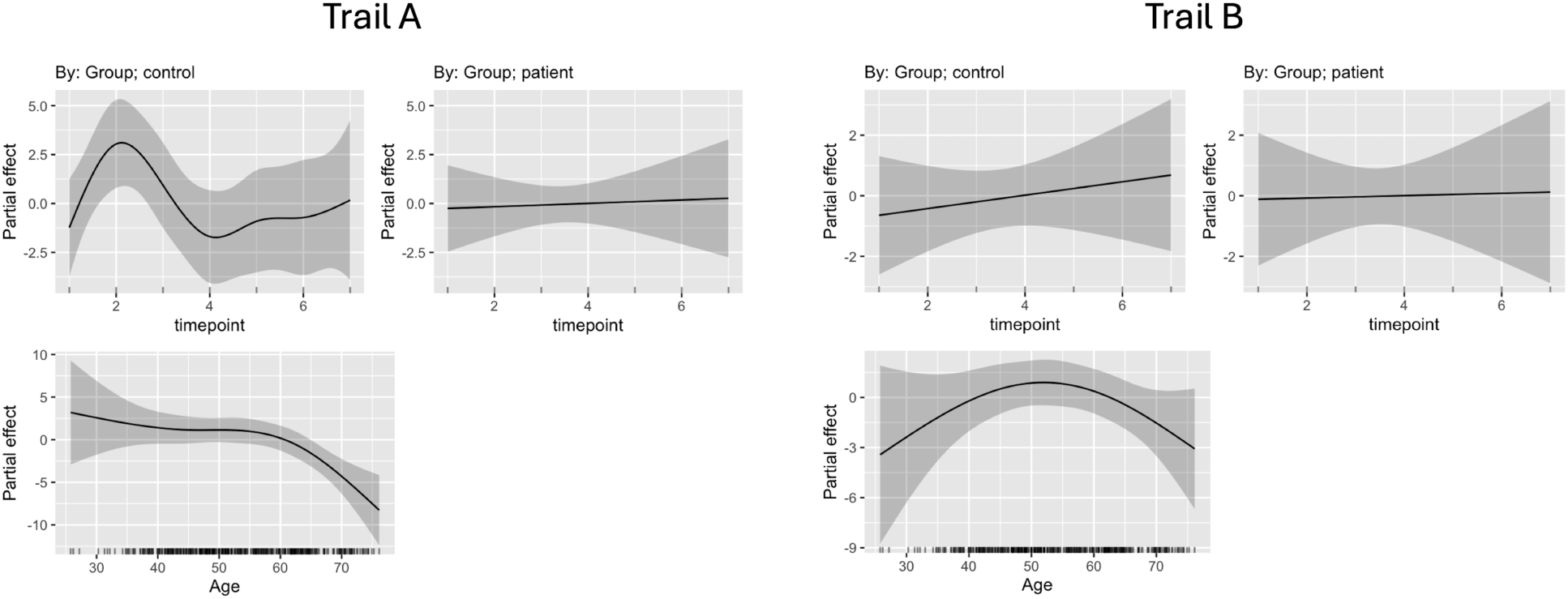
Longitudinal trajectories of cognitive performance. Patients demonstrated significantly lower scores on the Trails A (t = -4.29, p < 0.001) and B (t = -3.14, p = 0.002) tests of executive function compared to controls but trajectories of performance did not differ between groups. Plots show the partial effects controlling for all other predictors in the model. The solid line indicates the estimated smooth function for the generalized additive model with the shaded area representing the 95% confidence interval.

In patients, a model of Trails A performance that included gene expression and demographics (age and education) showed significantly higher goodness-of-fit compared to a demographics only model (χ² = 22.19, p = 0.003). *GRIN2A* (F = 4.39, p = 0.009) and age were significant predictors (F = 4.82, p = 0.003). The partial effects of *GRIN2A* expression and age on Trails A performance were nonlinear (Figure 3).

**Figure 3.**
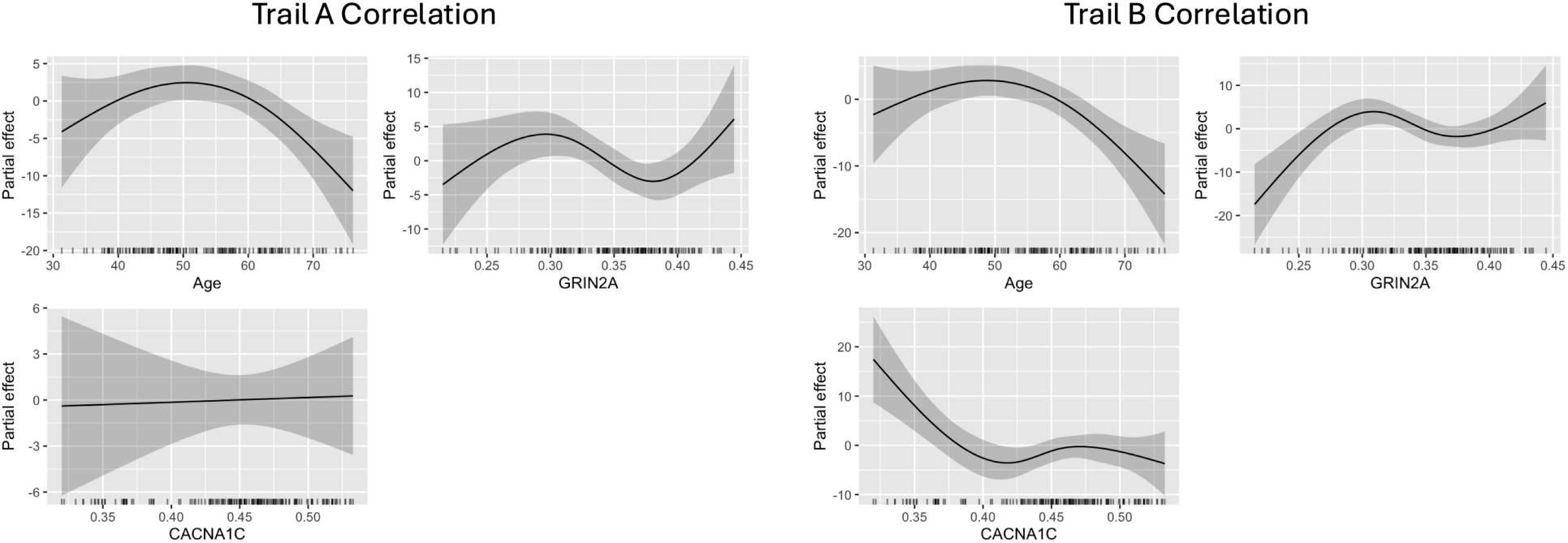
Predictors of cognitive performance in patients with breast cancer. In patients, *GRIN2A* expression (F = 4.39, p = 0.009) and age were significant predictors (F = 4.82, p = 0.003) of Trails A performance. *GRIN2A* (F = 6.25, p < 0.001), *CACNA1C* (F = 5.06, p = 0.002), age (F = 5.69, p = 0.001), and education (t = 4.20, p < 0.001) were significantly associated with Trails B performance. Plots show the partial effects controlling for all other predictors in the model. The solid line indicates the estimated smooth function for the generalized additive model with the shaded area representing the 95% confidence interval.

For Trails B, the model that included gene expression demonstrated significantly higher goodness-of-fit compared to a demographics only model (χ² = 37.38, p < 0.001). *GRIN2A* (F = 6.25, p < 0.001), *CACNA1C* (F = 5.06, p = 0.002), age (F = 5.69, p = 0.001), and education (t = 4.20, p < 0.001) were significantly associated with Trails B performance over time. The partial effects of age, *GRIN2A*, and *CACNA1C* were nonlinear (Figure 3).

## Discussion

*GRIN2A/B* encode NMDAR subunits which are ionotropic glutamate receptors critical for excitatory neurotransmission. NMDARs are unique from other glutamate receptors in several ways that impact cognitive functions. They allow passage of calcium in addition to sodium and potassium ions. Additionally, they are both ligand- and voltage-gated, requiring the coincidence of presynaptic glutamate and postsynaptic depolarization to pass calcium, which supports long-term synaptic information storage.[30] NMDARs have slower and longer-lasting amplitudes compared to other glutamate receptors, allowing sustained neural activity over time in the absence of sensory stimuli.[16, 31] These properties make NMDARs critical for executive skills, consistent with our findings that *GRIN2A* was significantly associated with Trails A/B performance. Trail Making Test performance is frequently impaired in patients with breast cancer.[32–34] Executive dysfunction is particularly problematic for cancer survivors given its strong association with treatment compliance, health behaviors, and fall risk, among others.[35–38]

*CACNA1C* was significantly associated with Trails B performance. Calcium signaling impacts neuronal excitability, neurotransmitter release, and gene expression in the brain. Both gain- and loss-of-function in calcium signaling can result in cognitive deficits.[39] Voltage-gated calcium channels, including CAv1.2, are involved in cell proliferation which makes them of interest for understanding cancer progression and identifying treatment targets. Several calcium channel encoding genes, including *CACNA1C*, have been shown to be highly expressed in various cancer tissue types, including breast cancer.[40] Mutations in *CACNA1C* have been associated with overall survival rate in certain cancers.[41, 42] Further study is required to determine if *CACNA1C* drivers of cancer pathophysiology are mechanistically linked to *CACNA1C* expression in the brain.

The partial associations between gene expression and cognitive performance were nonlinear, which is not surprising given that biological systems tend to be complex. Associations showed biphasic or inverted U-shaped relationships, which could suggest that low levels of gene expression may not be sufficient to influence downstream pathways, so cognition would not change until expression passed a certain point. Once above the threshold, effects may accelerate. At higher expression, the system may reach a plateau, so additional increases have diminishing or no effect. Certain levels of expression could be compensatory, while other levels of expression reflect dysregulation. There is likely an optimal range of gene expression in the brain that does not conform to traditional linear measurements.

Cancer treatments suppress neuroprogenitor cells while aging and inflammation disrupt mechanisms that protect neurons from the toxic effects of dysregulated calcium levels.[39, 43] This may result in increased neuron death (i.e., decreased gray matter), and thus, lower gene expression. Prior longitudinal studies of gray matter volume in patients with CRCI indicate acute atrophic changes followed by recovery.[44, 45] We observed that total gray matter volume showed an overall decreasing trend but was not significantly lower compared to controls. Prior studies involved a much shorter follow-up timeframe compared to ours (1-2 years vs. 10 years) but included regional analyses. We noted that gray matter volume contributed significantly to all three gene models. The partial relationship between gray matter and gene expression was nonlinear. Thus, gray matter changes may help explain differences in gene expression, but not sufficiently, or they may be driven by regional fluctuations that were not evaluated here.

*GRIN2B* expression did not differ between groups, nor did it change over time in either group. Although this gene was significantly and reliably expressed in gray matter, it was associated with the lowest variance explained (9.4% vs. 38%-48%). However, the lower expression of *GRIN2B* compared to *GRIN2A* is expected given that GluN2B receptors dominate during early development but are gradually replaced by GluN2A receptors with age.[46, 47] GluN2B receptors continue to play critical roles in synaptic plasticity and neurotransmission throughout the lifespan but have different trafficking and maintenance mechanisms in mature synapses compared to GluN2A.[48]

Pathological imbalances between GluN2A/B subunit interactions, localization, and trafficking are believed to play critical roles in several neurodegenerative conditions.[49] Functional synaptic NMDARs (highly enriched in GluN2A in mature neurons) support pro-survival and plasticity-related signaling pathways. When synaptic GluN2A is lost, the overall NMDAR activity becomes dominated by extrasynaptic NMDARs, which include GluN2B subunits. Extrasynaptic NMDARs are mechanistically linked to neuronal dysfunction and cell death pathways, often by suppressing the pro-survival signaling of their synaptic counterparts.[49, 50] Reduced *GRIN2A* expression may result in fewer available GluN2A subunits. Lower activity of pro-survival and plasticity-related signaling pathways results in negative feedback loops that suppress *GRIN2A* expression.

GluN2A/B subunits also interact with different scaffolding proteins, which determines their stability and location in the mature neuron. If a disease or injury affects the anchor proteins responsible for keeping a receptor at the synapse (e.g., PSD-95 binding sites), GluN2A would be the primary target because it has been trafficked in to define the mature synapse.[48, 51] GluN2B has a mechanism for internalization and as noted above, is largely extrasynaptic[52] and thus might be affected differently or less acutely by an impairment in the GluN2A-anchoring system. There is evidence showing that chemotherapies, including those used to treat breast cancer, functionally impair the PSD-95 complex and PSD-95 has been proposed as a mechanistic target for treating CRCI.[53, 54]

The reduced expression of *CACNA1C* in gray matter may be due to its regulatory role in the same activity-dependent and pro-survival gene networks as *GRIN2A*. Both *GRIN2A* and *CACNA1C* expression are positively regulated by cAMP response element-binding protein (CREB).[55] Healthy, synaptic activity activates CREB, which then promotes the transcription of genes necessary for long-term plasticity, maturation, and survival. A reduction in the efficiency of one channel often leads to a failure in the downstream signaling, which then triggers the suppressive feedback loop that also affects the other channel, leading to a convergent collapse of the cellular machinery responsible for generating and sensing pro-survival calcium signals.

In addition to supporting PSD-95 as a target for treating CRCI, our findings provide other translational insights for possibly addressing CRCI in the future. For example, neuron derived exosomes (NDEs) have the potential to act as intervention targets for neurodegeneration by delivering neuroprotective cargo such as microRNAs that promote repair.[56] Integrating transcriptomic data with exosomal microRNA cargo profiling would enable pathway-level inference, indicating whether NDE microRNAs regulate the same NMDA/calcium signaling genes disrupted in CRCI.

This study represents the first investigation of brain transcriptome changes associated with CRCI. Other innovative aspects of this research include the pre-surgical baseline and the approximately 10-year longitudinal span of follow-up assessments. However, there are also several limitations that should be considered. The sample became increasingly sparse across timepoints. To mitigate this, we used statistical models that borrow strength across adjacent observations, accommodate nonlinear trajectories, and include penalties to reduce overfitting in sparse regions. Nonetheless, reduced statistical power remains a concern, particularly for detecting subtle effects or interactions. Even though the AHBA transcriptomic atlas is the most comprehensive tool to date, it was developed based on only six donors. The atlas includes expression patterns for thousands of brain regions but does not cover the entire brain. Genes were selected based on specific hypotheses and with consideration of statistical power, but other genes may also be relevant for CRCI. Similarly, we chose to focus on specific neuropsychological tests, but other assessments may yield different results. We did not have sufficient samples to examine regional effects. Future research involving larger groups should explore whether variations in gene expression across different brain regions are associated with distinct patterns in cognitive performance.

Despite these limitations, our findings that expression of neurotransmission-related genes is reduced in patients with breast cancer align with prior evidence on the roles of these genes in brain function and neurodegeneration. These results provide novel insights into the potential molecular mechanisms underlying CRCI, addressing a critical gap in the current literature. Furthermore, this study establishes a strong foundation for ongoing and future investigations into the effects of cancer and its treatments on the brain transcriptome. Patients diagnosed with cancer experience cognitive deficits because of the tumor biology and the additive negative effects of cancer treatments on their cognition. These longitudinal data present a unique opportunity to understand the trajectory of CRCI long-term and its underlying molecular mechanisms. This research also offers broader insights into cognitive decline seen in other disorders where there is an interaction between gene expression, aging, disease pathophysiology, or stressors/injuries.

## Data Availability

All available data are contained in the manuscript

## Acknowledgements

The authors wish to express appreciation to the faculty and staff of the Richard M. Lucas Center for Imaging at Stanford University for assistance with neuroimaging acquisitions.

## Funding

This work was supported by research grants from the National Institutes of Health (1R01CA226080, 1R01CA172145, 2R01CA172145). The funder did not play a role in the design of the study; the collection, analysis, and interpretation of the data; the writing of the manuscript; and the decision to submit the manuscript for publication.

## Conflicts of Interest

None to report.

## Notes

### Competing Interest Statement

The authors have declared no competing interest.

### Funding Statement

This study was supported by research grants from the National Institutes of Health (1R01CA226080, 1R01CA172145, 2R01CA172145).

### Author Declarations

This study was approved by the Stanford University Institutional Review Board, was conducted in accordance with the ethical standards of the Declaration of Helsinki, and all participants provided written informed consent.

